# Expression of deubiquitinases in human gingiva and cultured human gingival fibroblasts

**DOI:** 10.1101/2021.01.26.21250532

**Authors:** Yong-Wei Fu, Hong-Zhi Xu

**Author notes:** Correspondence: Yong-Wei Fu, Department of Stomatology, The First People’s, Hospital of Lianyungang, 6 East Zhenhua Road, Haizhou District, Lianyungang 222061, Jiangsu, China.

## Abstract

**Background:** Although deubiquitinating enzymes (DUBs) such as CYLD, A20 and OTULIN are expressed in multiple tissues and thought to be linked with inflammatory diseases, their expression in periodontal tissues remains to be determined. This research was designed to assess the expression of CYLD, A20 and OTULIN in human gingiva, and to evaluate the regulation of these DUBs in human gingival fibroblasts (HGFs) upon different stimuli.

**Methods:** Immunohistochemistry assay was conducted to determine the expression of CYLD, A20 and OTULIN in human gingiva. Immunofluorescence assay was employed to observe the protein expression of CYLD, A20 and OTULIN in HGFs. RT-PCR and western blots were carried out to assess gene and protein expression changes of these DUBs in HGFs upon LPS or TNF-α.

**Results:** CYLD, A20 and OTULIN were found to be expressed in human gingiva and HGFs. Further, the expression of CYLD, A20 and OTULIN in HGFs exhibited distinct regulation by different stimuli.

**Conclusions:** Our findings suggest that CYLD, A20 and OTULIN might play a role in the progression of periodontitis.

## Background

Ubiquitin, a protein containing 76 amino acids, has an essential role in variety of biological processes [1-4]. Ubiquitination is a critical post-translational protein modification, performed by specific ubiquitin enzymes [5, 6]. The ubiquitin enzymes attach ubiquitin to target proteins and modulate the function of the target proteins. Attached ubiquitins can be removed by deubiquitinating enzymes (DUBs). Several DUBs including CYLD, A20 and OTULIN are reported to inhibit NF-κB activation and play an important role in modulating immunity and inflammation [7-9]. Furthermore, previous studies have shown that CYLD, A20 and OTULIN are all related to inflammatory diseases [10-13].

Periodontitis is an inflammatory disease of periodontal tissues which is caused by plaque biofilms [14]. In addition to being an important cause of tooth loss in adults, periodontitis is highly linked with multiple systemic diseases [15]. Recent work suggests that dysregulation of immune and inflammatory responses in the periodontium may be involved in the progression of periodontitis [16, 17]. Briefly, periodontal pathogenic bacteria and their toxic substances interact with host immune cells, result in immune and inflammatory responses in periodontal tissues, trigger the excessive release of inflammatory cytokines, and eventually cause periodontal tissue destruction [18]. Recent work demonstrates that regulating the immune responses may act as an adjunct to traditional periodontal therapy [19, 20].

NF-κB is an important regulator of immune and inflammatory responses [8, 21]. Upregulated NF-κB activity is reported in many inflammatory conditions such as inflammatory bowel disease, rheumatoid arthritis, and periodontitis [22, 23]. Evidence increasingly suggests that NF-κB activation may be involved in the progression of periodontitis [22, 24]. Periodontal pathogens may promote the induction of proinflammatory cytokines in periodontal tissues by upregulating NF-κB activation [18]. Considering the repressing effect of CYLD, A20 and OTULIN on NF-κB activation, it is reasonable to postulate that these DUBs may restrict periodontal inflammatory responses.

Although CYLD, A20 and OTULIN are expressed in multiple tissues, their expression in periodontal tissues remains to be elucidated. The objective of this research is to assess the expression of CYLD, A20 and OTULIN in human gingiva and to evaluate the regulation of these DUBs in HGFs upon different stimuli.

## Methods

### Antibodies and reagents

Rabbit anti-CYLD (catalog #ab137524), rabbit anti-A20 (catalog #ab92324) and rabbit anti-OTULIN (catalog #ab151117) were purchased from Abcam (Cambridge, UK). Normal rabbit IgG (catalog #A7016) were purchased from Beyotime Biotechnology (Shanghai, China). Alexa Fluor 568 goat anti-rabbit IgG (catalog #A11036) was supplied by Thermo Fisher Scientific (Rockford, IL, USA). Tubulin rabbit polyclonal antibody (catalog #10094-1-AP) and horseradish peroxidase-conjugated secondary antibody (catalog #SA00001-2) were supplied by Proteintech Group (Wuhan, China). Minimum essential medium alpha was supplied by Thermo Fisher Scientific (Suzhou, China). Penicillin-streptomycin solution (catalog #SV30010) was supplied by HyClone (Austria). Fetal bovine serum (catalog #VS500T) was supplied by Ausbian (Australia). Immunohistochemistry application solutions kit (catalog #13079) was supplied by Cell Signaling Technologies (Danvers, MA, USA). DAPI (catalog #R37606) and ProLong Diamond Antifade Mountant (catalog #P36965) were supplied by Molecular Probes (Eugene, OR, USA). TNF-α (catalog #01375) was supplied by R&D Systems (Minneapolis, MN, USA). LPS (catalog #tlrl-pglps) was supplied by InvivoGen (San Diego, USA). RNA extraction kit (catalog #R6834-01) was from OMEGA Bio-Tek (Norcross, GA, USA). PrimeScript RT Master Mix kit (catalog #RR036A) and SYBR Premix Ex Taq □ kit (catalog #RR820A) were from Takara Bio Inc. (Otsu, Japan). BCA Protein Assay Kit (catalog #23227) was from Pierce Biotechnology (Rockford, IL, USA).

### Gingival samples

Human gingival biopsy samples were acquired from 12 individuals (5 male and 7 female, average age of 28.7 y) undergoing crown-lengthening surgery. All individuals were selected from the First People’s Hospital of Lianyungang between March 2019 and August 2019. Informed consent forms were assigned by all individuals before they were enrolled in the research. This study was approved by The Ethics Committee of Nanjing Medical University (2018389). All experiments on human subjects were conducted in accordance with the 1964 Helsinki declaration and its later amendments or comparable ethical standards.

### Immunohistochemistry assay

Gingival samples were fixed in 4% paraformaldehyde for 24 h at 4□. Then these samples were processed to paraffin blocks and sectioned at 5 µm. Immunohistochemical study was undertaken as previously described using a commercial immunohistochemistry kit [4]. Anti-CYLD (1:400), anti-A20 (1:400) and anti-OTULIN (1:400,) antibodies were applied in this study. Nonimmunized IgG (1:400, Beyotime Biotechnology) was used as negative controls in the present study. Slices were photographed with a Leica DM4000 B microscope.

### Cell culture

HGFs were isolated from human gingival tissues and cultured with an explant culture method. Briefly, gingival tissues were acquired from three healthy individuals (2 females and 1 male, aged 11-14 y) experiencing first premolar extractions for orthodontic therapy. The extracted teeth were placed in phosphate buffered saline (PBS) and immediately transferred to the laboratory. Then the gingival tissues attached to the cervical region of the extracted teeth were removed and cut into small pieces in the biosafety cabinet. Afterwards, the tissue pieces were transferred to 25 cm^2^ culture flasks and cultured in minimum essential medium alpha supplemented with 15% fetal bovine serum. HGFs at passages 3 to 6 were used in this study.

### Immunofluorescence staining

For Immunofluorescence Staining, HGFs were seed in 24-well culture plates with a coverslip at the bottom in a density of 2.5×10^4^cells per well. After 24 h, HGFs were fixed with 4% paraformaldehyde in PBS for 15 min, permeabilized with 0.2% Triton X-100 in PBS for 5 min, blocked with 1% BSA in PBS for 1 h at room temperature (RT). Then HGFs were incubated with primary antibodies against CYLD (1:200), A20 (1:200) and OTULIN (1:200) overnight at 4□. Afterwards, HGFs were stained with Alexa Fluor 568-conjugated secondary antibody (4 μg/mL) for 50 min and then with DAPI for 6 min at RT. Each of the above-mentioned steps was followed by three 5-min washes in PBS. Finally, coverslips were mounted using ProLong Diamond Antifade Mountant and data were obtained by using a Leica DM4000 B microscope.

### Cell stimulation

HGFs were seed in 6-well culture plates and cultured for 24 h. After washing twice with PBS, HGFs were starved for 12 h in α-MEM without foetal bovine serum. Then HGFs were stimulated with 1 μg/ml LPS or 10 ng/ml TNF-α as indicated.

### RNA isolation and reverse transcription polymerase chain reaction (RT-PCR)

Total RNA was extracted from treated HGFs with an RNA extraction kit per the supplier’s protocol. cDNA synthesis was undertaken using a PrimeScript RT Master Mix kit. Then RT-PCR was carried out using a SYBR Premix Ex Taq □ kit on a 7300 Real Time PCR System. Each sample was run in triplicate. Results were normalized to GAPDH. Primers were: CYLD, 5□-ACGCCACAATCTTCATCACACT-3’ (forward:) and 5□-AGGTCGTGGTCAAGGTTTCACT-3’ (reverse); ; *A20*, 5’- ATGCACCGATACACACTGGA-3’ (forward) and 5’- GGATGATCTCCCGAAACTGA-3’ (reverse); *OTULIN*, 5’- AAAGAGGGGCATCAGAACCG-3’ (forward) and 5’- GGCCCTCAGTGCACAGTAAT-3’ (reverse); and GAPDH, 5’- TGCACCACCAACTGCTTAGC-3’ (forward) and 5’-GGCATGGA- CTGTGGTCATGAG-3’ (reverse).

### Protein extraction and western blotting

For western blotting, pre-treated HGFs were lysed using RIPA lysis buffer containing protease inhibitors. The protein concentrations were determined with a BCA Protein Assay Kit. Then, the protein extracts were subjected to western blotting with antibodies to CYLD (1:1000), A20 (1:1000), OTULIN (1:400), and Tubulin (1:1000). Horseradish peroxidase-conjugated secondary antibody (1:8000) was applied. Densitometry quantification of the protein bands was processed with ImageJ software.

### Statistical analysis

Data are expressed as the mean ±SD. ANOVA followed by Dunnett’s multiple comparison test was used to determine the significance of differences. GraphPad Prism 5 was used to perform statistical analyses. Differences were considered to be significant when *P* <0.05.

## Results

### Expression of deubiquitinases in human gingival tissues

To determine the expression of CYLD, A20 and OTULIN in human gingiva, we performed immunohistochemistry staining on gingival tissue samples from 12 individuals. Positive staining cells for CYLD, A20and OTULIN were observed both in the epithelium and in the connective tissue (Fig. 1). Further, all of the above deubiquitinases were mainly expressed in epithelial cells.

**Fig. 1.**
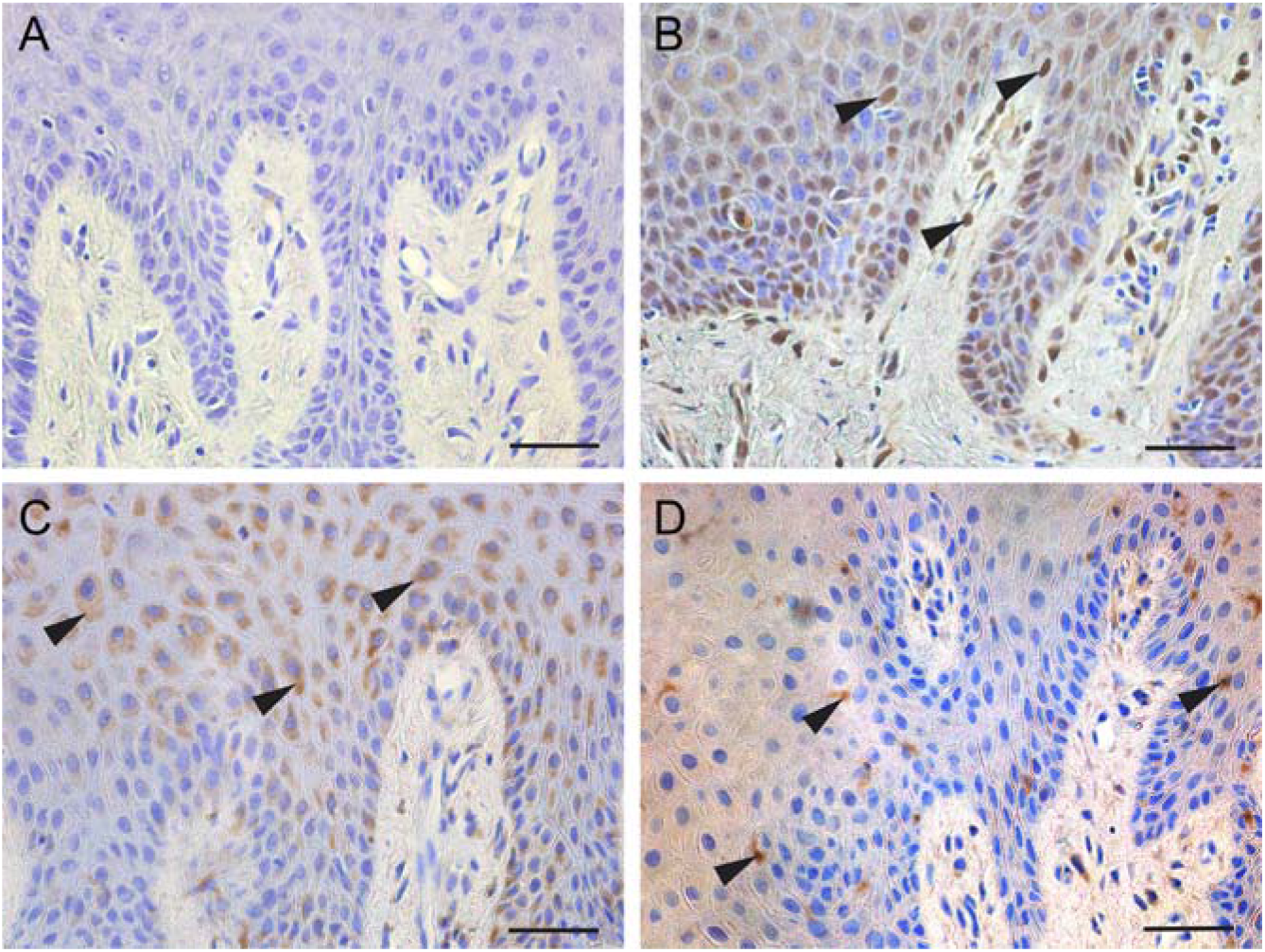
Immunohistochemistry for CYLD, A20 and OTULIN on sections from gingival tissue. **a** The negative control of immunohistochemistry. **b** CYLD staining in gingival tissue. Arrows show positive staining for CYLD. **c** A20 staining in gingival tissue. Arrows show positive staining for A20. **d** OTULIN staining in gingival tissue. Arrows show positive staining for OTULIN. Scale bar, 50 μm

### Expression of deubiquitinases in human gingival fibroblasts

Immunofluorescence was performed to assess the expression of deubiquitinases in HGFs. As expected, HGFs displayed positive staining for CYLD, A20 and OTULIN. (Fig. 2).

**Fig. 2.**
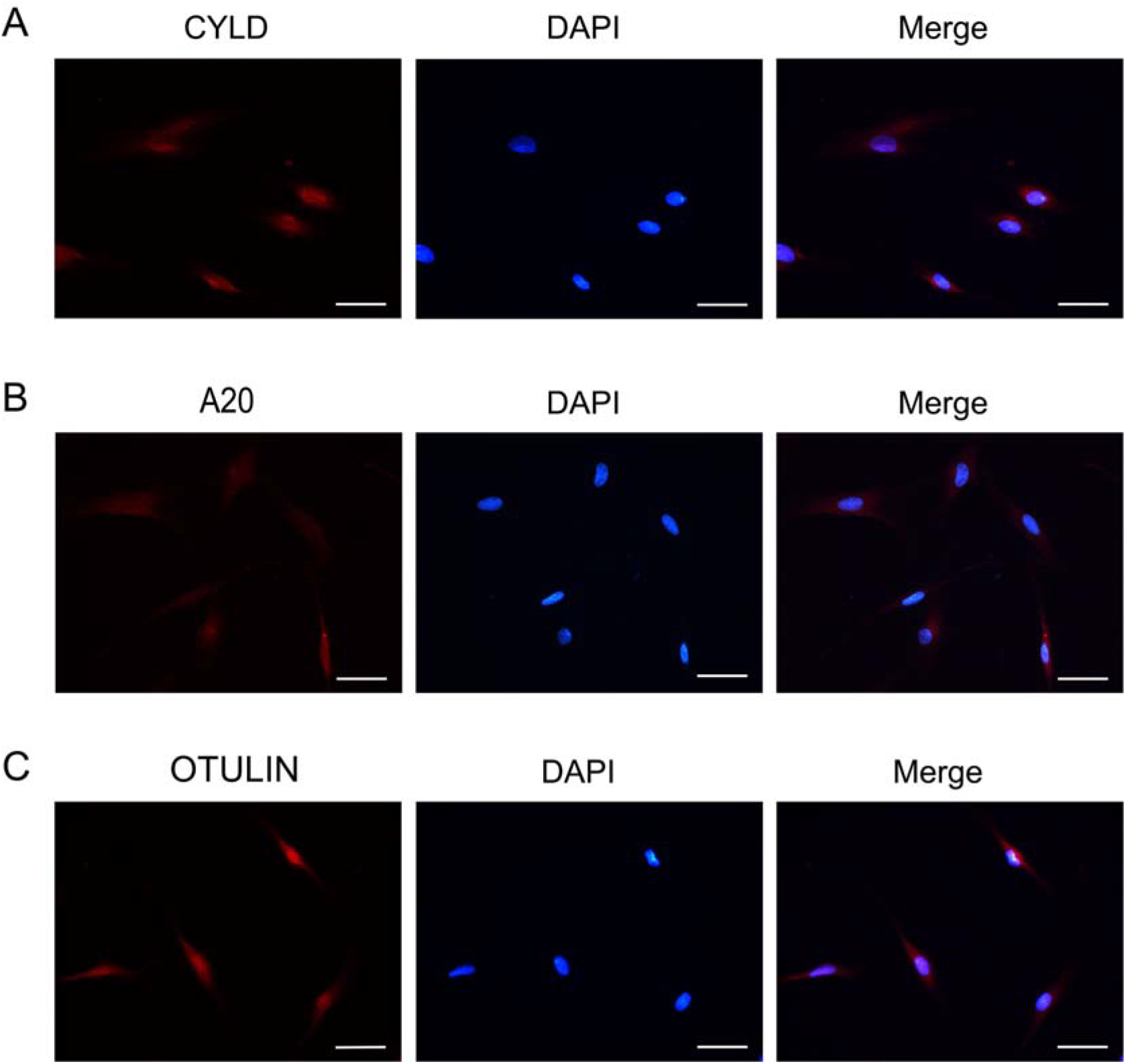
Immunofluorescence staining for CYLD (**a**), A20 (**b**) and OTULIN (**c**) in HGFs. Scale bar, 50 μm

### Regulation of deubiquitinases by different stimuli

To examine the changes in the expression levels of deubiquitinases in LPS-or TNF-α-treated HGFs, RT-PCR and western blotting were performed. Results demonstrated that the expression of CYLD, A20 and OTULIN in HGFs exhibits distinct regulation upon LPS or TNF-α (Fig. 3 and Fig. 4).

**Fig. 3.**
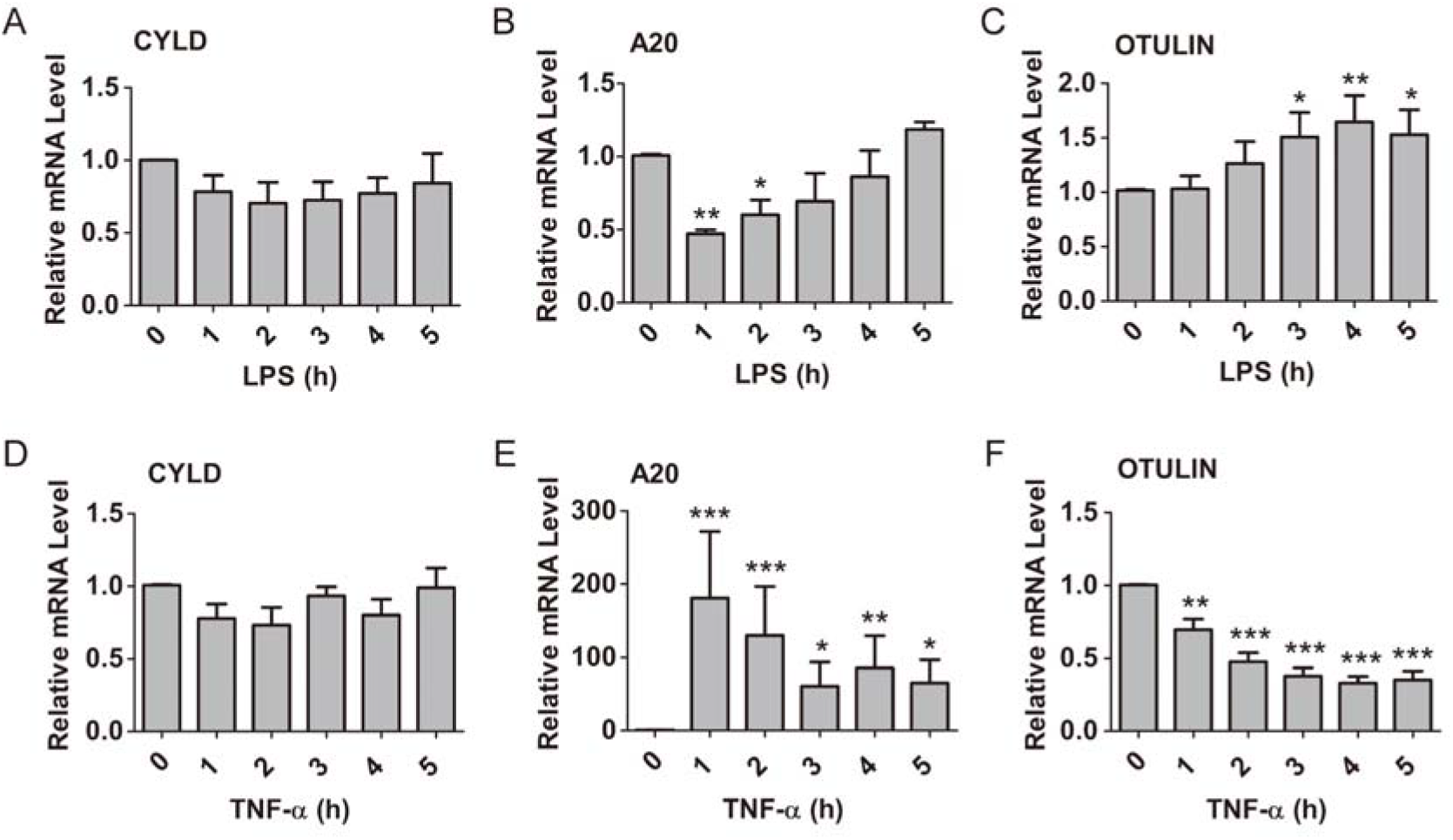
mRNA levels of CYLD, A20 and OTULIN in HGFs pretreated with LPS (**a, b, c**) or TNF-□ (**d, e, f**) for the indicated periods of time. ^⍰^*P*<0.05, ^⍰⍰^*P*<0.01, ^⍰⍰⍰^*P*<0.001, 0 h versus 1h, 2h, 3h, 4h or 5 h, one-way ANOVA with Dunnett’s multiple comparison test. Data presented are from three independent experiments

**Fig. 4.**
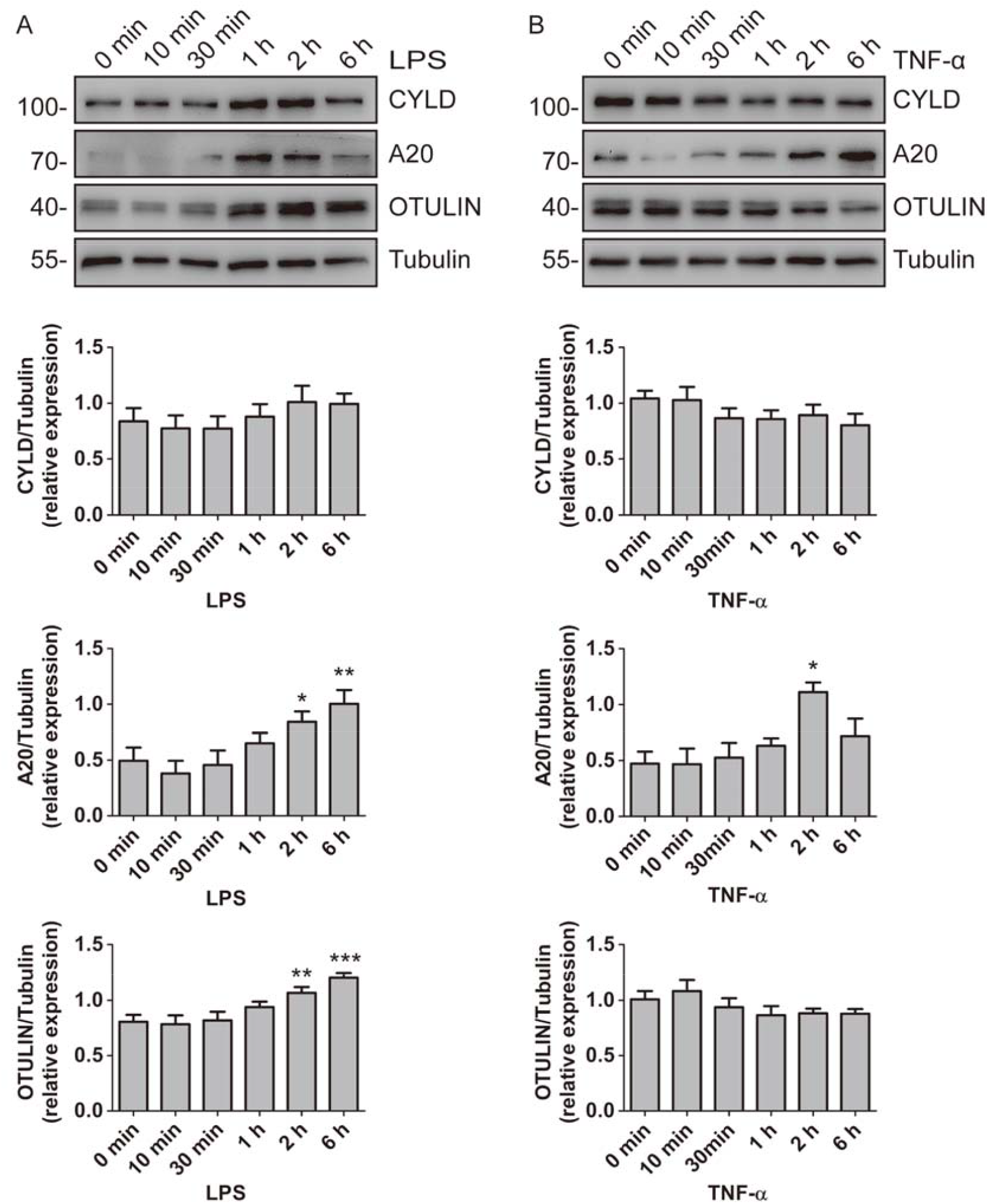
Protein expression of CYLD, A20 and OTULIN in HGFs pretreated with LPS (**a**) or TNF-α (**b**) for the indicated periods of time. Below, quantification of the band intensity results, presented relative to Tubulin. ^⍰^*P*<0.05, ^⍰⍰^*P*<0.01, ^⍰⍰⍰^*P*<0.001, 0 h versus 2h or 6h, one-way ANOVA with Dunnett’s multiple comparison test. Data presented are from three independent experiments

The RT-PCR results demonstrated that the CYLD mRNA level had no obvious change after treatment with LPS or TNF-α (Fig. 3a and 3d). Intriguingly, the mRNA expression of A20 was decreased 1 h after treatment with LPS, and returned to the baseline level after 5 h (Fig. 3b). As expected, the mRNA expression of A20 was rapidly upregulated in response to TNF-α (Fig. 3e). In addition, LPS led to an increase in OTULIN mRNA expression after 3 h (Fig. 3c). In contrast, TNF-α resulted in a decrease in the mRNA expression of OTULIN after 1 h (Fig. 3f).

At the protein level, CYLD expression had no significant change upon LPS or TNF-α treatment (Fig. 4a and 4b). In addition, the protein expression of A20 markedly increased after treatment with LPS or TNF-α (Fig. 4a and 4b). Furthermore, LPS also led to the increase of OTULIN protein expression (Fig. 4a). However, TNF-α treatment did not affect the protein expression of OTULIN (Fig. 4b).

## Discussion

It is known that DUBs such as CYLD, A20 and OTULIN can inhibit inflammatory responses by repressing NF-κB and other signaling pathways [10, 11, 25, 26]. In the present study, both biopsies of human gingiva and HGFs show the expression of CYLD, A20 and OTULIN. Furthermore, the expression of CYLD, A20 and OTULIN in HGFs exhibits distinct regulation by different stimuli. Our findings thus imply that CYLD, A20 and OTULIN may play a part in periodontal inflammatory responses.

Previous studies demonstrated that CYLD, A20 and OTULIN are all critical players in suppressing inflammation and immunity [7, 9, 26]. The deficiency of these DUBs may lead to constitutive NF-κB responses, enhance NF-κB-dependent gene expression, and result in excessive production of proinflammatory cytokines [5, 8, 9]. In our present study, we characterized the expression of CYLD, A20 and OTULIN in human gingiva and HGFs. In line with our findings, a previous study also confirmed A20 expression in human gingival tissue [27]. Given the role of these DUBs in inhibiting inflammation, we postulate that these DUBs may be involved in the development of periodontitis.

It is known that CYLD expression can be upregulated by many stimuli. In the present study, we found that both LPS and TNF-α did not increase the mRNA level of CYLD. Similarly, western blotting data also showed that the protein level of CYLD had no obvious change in the presence of LPS or TNF-α. These findings are in line with our previous study [4]. In contrast, several early reports indicated that CYLD expression was upregulated by LPS or TNF-α treatment [28-30]. The potential mechanisms for this discrepancy remain unknown, but probably due to cell-type-specific differences.

A20, a key regulator of inflammation, is expressed at a low level in most cell types and can be induced by various stimuli including TNF-α, LPS, and IL-1 [9]. In this study, the mRNA level of A20 was markedly upregulated by TNF-α. In contrast, LPS treatment led to a decrease in A20 mRNA expression. As expected, both LPS and TNF-α led to the increase of A20 protein expression. We found a lack of concordance between the protein and mRNA levels of A20 in HGFs upon LPS treatment. This result was unexpected but interesting. Many prior works have demonstrated that the expression levels of mRNA and protein are often poorly correlated [31-33]. This discrepancy may be attributed to other types of regulation, such as post-transcriptional processing and protein degradation [31, 32]. In addition, several recent studies indicated that A20 may act as a critical regulator in the pathogenesis of periodontitis [27, 34-36].

OTULIN was identified as the deubiquitinase that specifically hydrolyzes linear polyubiquitin chains and suppresses NF-κB activation [37]. Recent work suggested that OTULIN can limit inflammation by deubiquitinating LUBAC [10]. However, the regulation of OTULIN is yet to be determined. A recent work showed that porcine reproductive and respiratory syndrome virus infection upregulates the expression of OTULIN [38]. In our current study, LPS resulted in the increase of the mRNA and protein expression of OTULIN. In contrast, the OTULIN mRNA level was decreased upon TNF-α stimulation. Unexpectedly, TNF-α treatment did not change the protein expression of OTULIN. Further studies need to be carried out to investigate the precise mechanism of OTULIN regulation.

## Conclusions

In summary, we show that the DUBs CYLD, A20 and OTULIN are expressed in human gingival tissues and are distinctly regulated by different stimuli. Given the role of these DUBs in limiting inflammation, our findings suggest that CYLD, A20 and OTULIN might be involved in the pathogenesis of periodontitis and might act as potential therapeutic targets to treat periodontitis. Therefore, the exact effects of these DUBs on periodontal inflammatory responses require further investigation.

## Data Availability

The data set supporting the results of this article are available from the corresponding author.

## Abbreviations

DUBs: Deubiquitinating enzymes
HGFs: Human gingival fibroblasts
PBS: Phosphate buffered saline
RT-PCR: Reverse transcription polymerase chain reaction

## Acknowledgments

We are grateful to Yan-Hui Li for her critical reading of the manuscript.

## Authors’ Contributions

Yong-Wei Fu is the major person to complete this study. Hong-Zhi Xu is the major person to write the paper.

## Funding

This work was supported by Lianyungang Health Development Project with Science and Education (2017-26).

## Availability of data and materials

The datasets used in the current study are available from the corresponding author on reasonable request.

## Ethics approval and consent to participate

This study was approved by The Ethics Committee of Nanjing Medical University (2018389). Informed consent forms were assigned by all individuals before they were enrolled in the research.

## Consent for publication

Not applicable.

## Competing interests

The authors declare no conflicts of interest.

## Notes

### Competing Interest Statement

The authors have declared no competing interest.

### Author Declarations

This study was approved by The Ethics Committee of Nanjing Medical University (2018389)

